# Population-Level Distribution of PRISQ Scores among adults accessing primary care services in the state of Qatar

**DOI:** 10.64898/2026.03.26.26349362

**Authors:** Dana Bilal El Kaissi, Mohamed Ahmed Syed, Muslim Abbas Syed

**Affiliations:** Clinical Research-Primary Health Care Corporation Qatar, Doha, Qatar; Department of Clinical Research -Primary Health Care Corporation Qatar, Doha, Qatar

**Author notes:** **(Corresponding author)** Contact: 0097471234476, Personal. Contributions: Literature review, data analysis and interpretation, drafting of manuscript, review of manuscript. Contributions: Conception of idea, literature review, data analysis and interpretation, drafting of manuscript, review of manuscript.

**Keywords:** Prediabetes risk, PRISQ, Primary health care, Cardiometabolic risk, Qatar

## Abstract

**Background:** Prediabetes is a critical intermediate stage in the development of type 2 diabetes mellitus and is increasingly prevalent in the Eastern Mediterranean Region. In Qatar, high levels of metabolic and lifestyle-related risk factors underscore the need for scalable, non-invasive risk stratification tools within primary care. The Prediabetes Risk Score in Qatar (PRISQ) was developed as a population-specific screening tool; however, its distribution and associated risk patterns within national primary care settings remain insufficiently characterized. This study aimed to assess the population-level distribution of PRISQ scores among adults attending primary care in Qatar and to identify key sociodemographic and clinical correlates of elevated prediabetes risk.

**Methods:** A cross-sectional analysis was conducted among adults (≥18 years) registered with the Primary Health Care Corporation (PHCC), using data derived from the HEALTHSIGHT study. PRISQ scores were calculated based on five non-invasive clinical parameters: age, sex, body mass index, waist circumference, and blood pressure. Participants were categorized into low, moderate, and high-risk groups using established PRISQ cut-offs. Descriptive analyses summarized risk distributions, and multivariable linear regression was used to identify independent predictors of PRISQ scores.

**Results:** Among 1,116 participants included in the final analysis, the mean PRISQ score was 26.5 ± 11.0. Nearly half of the study population (47.7%) was classified as high risk for prediabetes, while 34.4% and 17.9% were categorized as moderate and low risk, respectively. Increasing age was the strongest contributor to higher PRISQ scores, followed by body mass index, waist circumference, and blood pressure (all p < 0.001). High-risk individuals were more frequently male, older, overweight or obese, and long-term residents of Qatar, with variation across nationality groups.

**Conclusions:** A substantial proportion of adults attending primary care in Qatar are at high predicted risk for prediabetes. These findings support the utility of PRISQ as a risk stratification and engagement tool in primary care to guide early lifestyle counselling and targeted preventive interventions. Longitudinal studies are needed to assess progression to dysglycemia and to further refine risk-based screening strategies.

## Introduction

Diabetes mellitus represents a growing global health crisis. Type 2 diabetes mellitus (T2DM), which constitutes nearly 90% of all diagnosed cases, contributes substantially to preventable morbidity and premature mortality, particularly when diagnosis or treatment is delayed [1, 2]. Prediabetes, an intermediate metabolic state characterized by glycemic dysregulation that does not yet meet the diagnostic threshold for T2DM, represents a critical juncture for prevention, as individuals in this state exhibit elevated metabolic and hemodynamic abnormalities including higher blood pressure levels[3]. With more than 400 million individuals affected globally, prediabetes constitutes a major public health concern, especially in low- and middle-income countries where its prevalence continues to rise[4].

The Middle East is recognized as a high-burden region for both prediabetes and diabetes, with countries in the Gulf Cooperation Council (GCC) including Saudi Arabia, Kuwait, and the United Arab Emirates—reporting prevalence estimates ranging between 20% and 40%[5-7]. In Qatar, a national survey reported an 11.9% prevalence of prediabetes among adults aged 18–64 years[8]. More recently, a screening-based study found that approximately 21% of Qataris were at high risk for developing prediabetes, underscoring the need for systematic risk assessment to guide early intervention[9].

Beyond its role as a precursor to T2DM, prediabetes is independently associated with increased risk of cardiovascular disease (CVD), one of the leading causes of mortality worldwide. Individuals with prediabetes frequently exhibit elevated blood pressure, dyslipidemia, and other cardiometabolic disturbances, reflecting underlying insulin resistance and impaired β-cell function[10]. Importantly, evidence suggests that timely identification and intervention— through lifestyle modification or pharmacologic therapy—can delay or even reverse progression to diabetes[11]. Conversely, if left unrecognized, 5–10% of individuals with prediabetes progress to T2DM annually[12]. These findings highlight the urgency of improving early detection strategies to mitigate downstream health and economic consequences[9].

Although laboratory-based diagnostic tools such as fasting plasma glucose (FPG), hemoglobin A1c (HbA1c), and oral glucose tolerance testing (OGTT) provide accurate identification of prediabetes, they remain invasive, costly, and logistically impractical for large-scale population screening [13]. This challenge has prompted interest in non-invasive, cost-efficient risk assessment tools suitable for broader deployment[14]. Established instruments such as the Finnish Diabetes Risk Score (FINDRISC) and the American Diabetes Association (ADA) risk score have demonstrated strong predictive value in their respective populations[15, 16]. However, their performance in the Middle Eastern context is limited by demographic, genetic, sociocultural, and lifestyle differences, as well as a lack of region-specific validation.

To address this gap, the Prediabetes Risk Score in Qatar (PRISQ) was developed using data from the Qatar Biobank cohort. Designed to reflect the demographic, genetic, and lifestyle characteristics of Qatar’s population, PRISQ incorporates five routinely collected, non-invasive clinical variables—age, sex, BMI, waist circumference, and blood pressure—and generates a total risk score ranging from 0 to 45[13]. In its derivation study, PRISQ demonstrated strong diagnostic performance, including a sensitivity of 86.2% and an AUC of 80% at a cut-off score of 16[13]. Given its simplicity and suitability for resource-limited settings, PRISQ holds promise for use across Middle Eastern populations with similar demographic and lifestyle characteristics[17].

Despite the high burden of Type 2 diabetes mellitus (T2DM) and prediabetes across the Gulf Cooperation Council (GCC), PRISQ has not been evaluated within national healthcare systems [18-20]. In Qatar, its score distribution and demographic and clinical correlates remain unexamined within the Primary Health Care Corporation (PHCC), the country’s main primary care provider. This gap underscores the need for context-specific validation of non-invasive screening tools to support early detection and intervention in primary care.

This study evaluates prediabetes risk in Qatar’s primary care setting using the PRISQ tool by assessing score distribution, established risk strata, and associations with key sociodemographic factors among PHCC attendees. The findings aim to inform the potential integration of PRISQ into routine primary care to support early risk identification, targeted prevention, and efforts to reduce the national diabetes burden.

## Methodology

### Study Design and Setting

A cross-sectional study was designed to assess the risk of prediabetes among adults attending primary care services in Qatar using the Prediabetes Risk Score Questionnaire (PRISQ) and to examine the association between PRISQ scores and sociodemographic characteristics of primary care attendees. The study was conducted in accordance with a previously published study protocol[21].

The study was implemented within the Primary Health Care Corporation (PHCC), the largest publicly funded primary care provider in Qatar. PHCC operates a nationwide network of 31 health centers and provides comprehensive primary care services to the population. In 2024, approximately 1.87 million individuals were registered with PHCC, making it an appropriate and representative setting for population-based risk assessment in the primary care context.

### Study Population and Sampling

The study population comprised individuals aged 18 years and older who were registered with PHCC, possessed a valid health card, and had available contact information recorded in PHCC’s electronic medical records (EMR) system. Individuals who were pregnant, as well as those with communication, logistical, or clinical limitations that could interfere with participation or data collection (including, but not limited to, bleeding disorders or mental disabilities), were excluded from the study.

A stratified random sampling approach was employed to enhance representativeness of the PHCC-registered population. The sampling frame was stratified according to age group (18–29, 30–39, 40–49, 50–59, and ≥60 years), sex (male and female), and nationality (Qatari, North African, Southeast Asian, Southern Asian, Western Asian, and other nationalities), as detailed in Table S2 (Supplementary Material). Each stratum was assigned a target of 100 participants, resulting in an intended total sample size of 6,000 individuals[21].

To account for an anticipated response rate of approximately 10%, invitations were distributed to 60,000 randomly selected eligible individuals to ensure attainment of the minimum desired sample size.

### Data Collection Procedures

Eligible individuals were invited to participate in the study via Short Message Service (SMS). The invitation message described the objectives and expected outcomes of the study and included a digital link to an online form through which recipients could express interest in participation. Individuals who consented electronically were subsequently invited to attend an in-person appointment at their nearest PHCC health center.

At the health centers, trained data collectors conducted participant visits following standardized procedures. Written informed consent was obtained from all participants prior to data collection. Data collection activities included administration of a structured questionnaire, collection of physical measurements, and venous blood sampling, as described in the study protocol[21]. In addition, data related to participants’ health service utilization over a one-year period were extracted from the PHCC EMR system.

Collected data were organized into two main domains: PRISQ-related metrics and sociodemographic variables. PRISQ metrics included age, sex, body mass index (BMI), waist circumference, and blood pressure. Sociodemographic variables comprised nationality, educational attainment, household income, duration of residence in Qatar, and self-reported physical activity. These variables were obtained from participant questionnaires and supplemented with relevant information extracted from EMR records.

### Calculation of PRISQ Scores and Risk Classification

PRISQ scores were calculated in accordance with previously published methodology (Abbas et al., 2021). Predefined scores were assigned to each of the five PRISQ components (age, sex, BMI, waist circumference, and blood pressure), as outlined in Table 1. The individual component scores were summed up to generate a total PRISQ score ranging from 0 to 45.

**Table 1.**
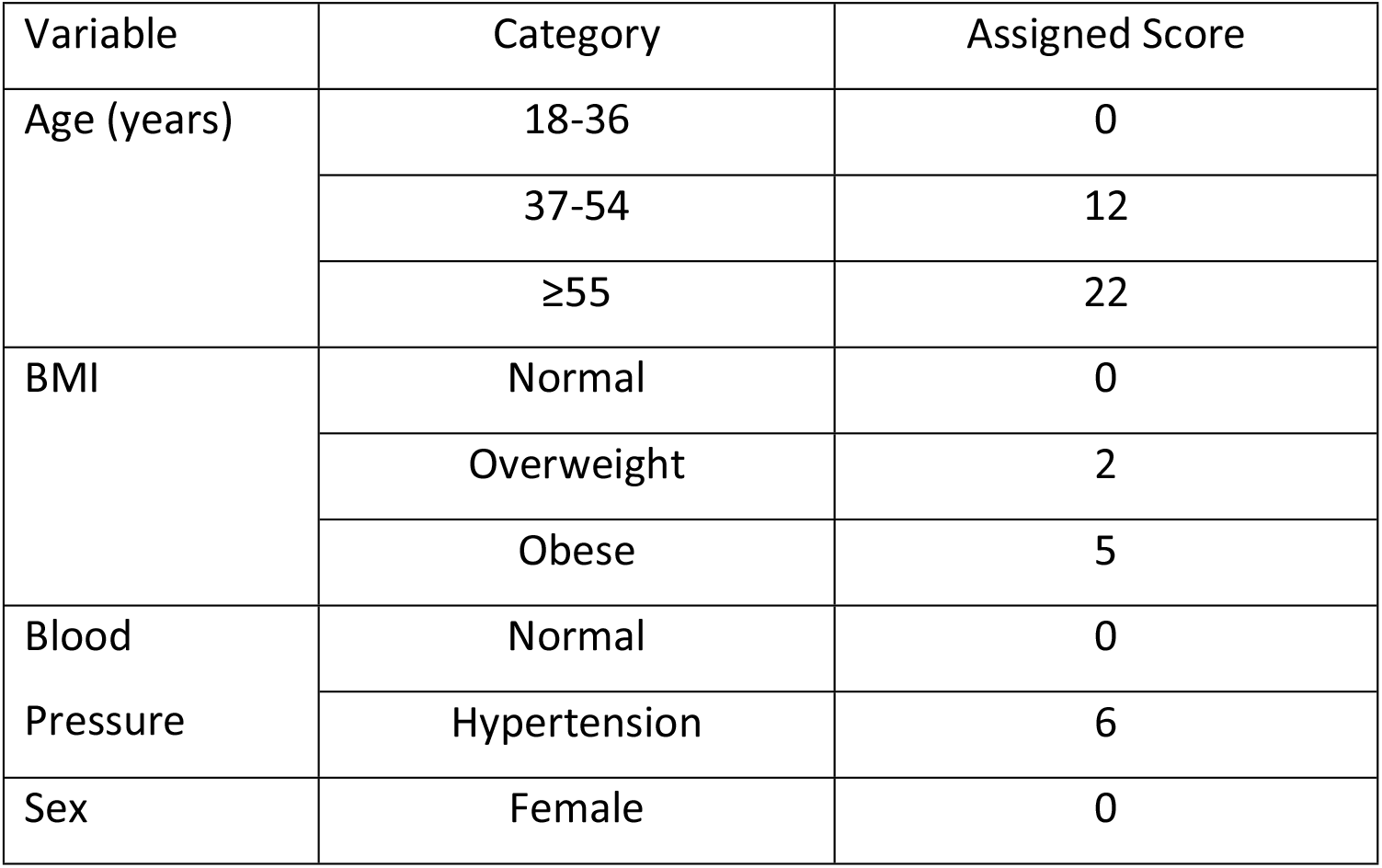
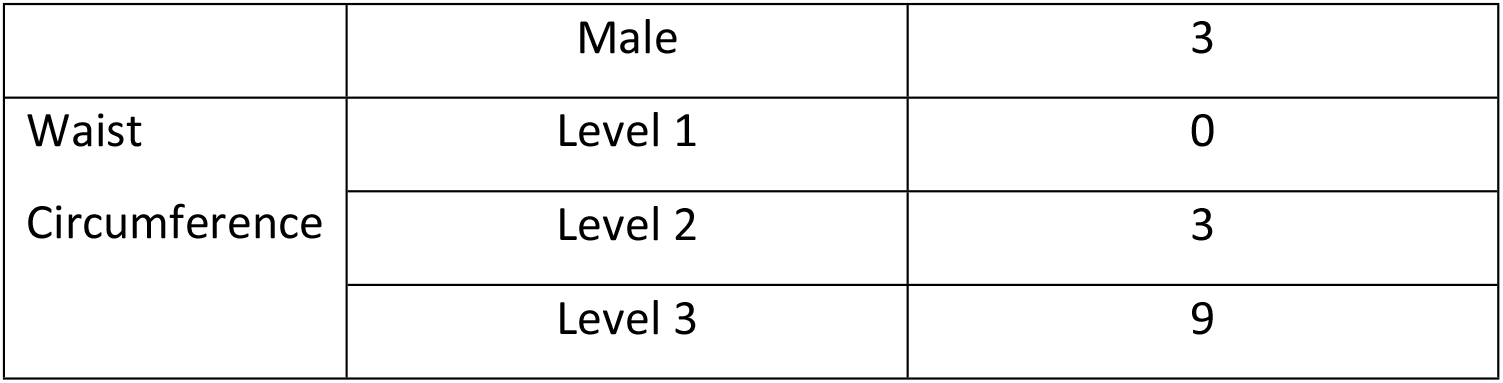
PRISQ Scoring System (adapted from [13].

Based on established cut-off values reported in earlier studies, participants were categorized into three prediabetes risk groups: low risk (PRISQ score ≤16), moderate risk (PRISQ score 17–27), and high risk (PRISQ score >27).

### Data Analysis

Data was cleaned, categorized, and analyzed using Microsoft Excel (Microsoft Corporation, Redmond, WA, USA) with its built-in statistical functions to facilitate data management and accessibility. Descriptive statistical analyses were initially performed to summarize the sociodemographic characteristics of the study population and to describe the distribution of PRISQ scores across predefined risk categories. Continuous variables were summarized using appropriate measures of central tendency and dispersion, while categorical variables were presented as frequencies and proportions.

To examine the associations between PRISQ risk categories and sociodemographic and clinical factors, multiple linear regression models were employed. PRISQ scores were treated as the dependent variable, and relevant sociodemographic and clinical variables were included as independent predictors. Model assumptions were assessed prior to analysis. Statistical significance was defined as a priori as a two-sided p-value < 0.05.

## Results

### Baseline Characteristics of the Study Population

A total of 1,960 individuals aged between 18 and 90 years initially responded and had their clinical and sociodemographic information extracted. Of these, 844 individuals were excluded due to missing data, resulting in a final analytic sample of 1,116 participants included in the study.

The final study population comprised a slightly higher proportion of males (54.3%, *n* = 606) compared with females (45.7%, *n* = 510). The mean age of participants was 47.8 years (±12.9 standard deviation [SD]). Overall, the clinical profile of the cohort was indicative of an overweight population, with a mean body mass index (BMI) of 29.7 ± 5.7 kg/m^2^. The average systolic blood pressure was 124.5 ± 16.4 mmHg, while the mean diastolic blood pressure was 77.3 ± 8.9 mmHg.

Assessment of prediabetes risk using the Prediabetes Risk Score Questionnaire (PRISQ) demonstrated that the study population was predominantly skewed toward a moderate-to-high risk profile, with a mean PRISQ score of 26.5 ± 11.0. Detailed baseline sociodemographic and clinical characteristics of the study participants are presented in Table 2.

**Table 2.**
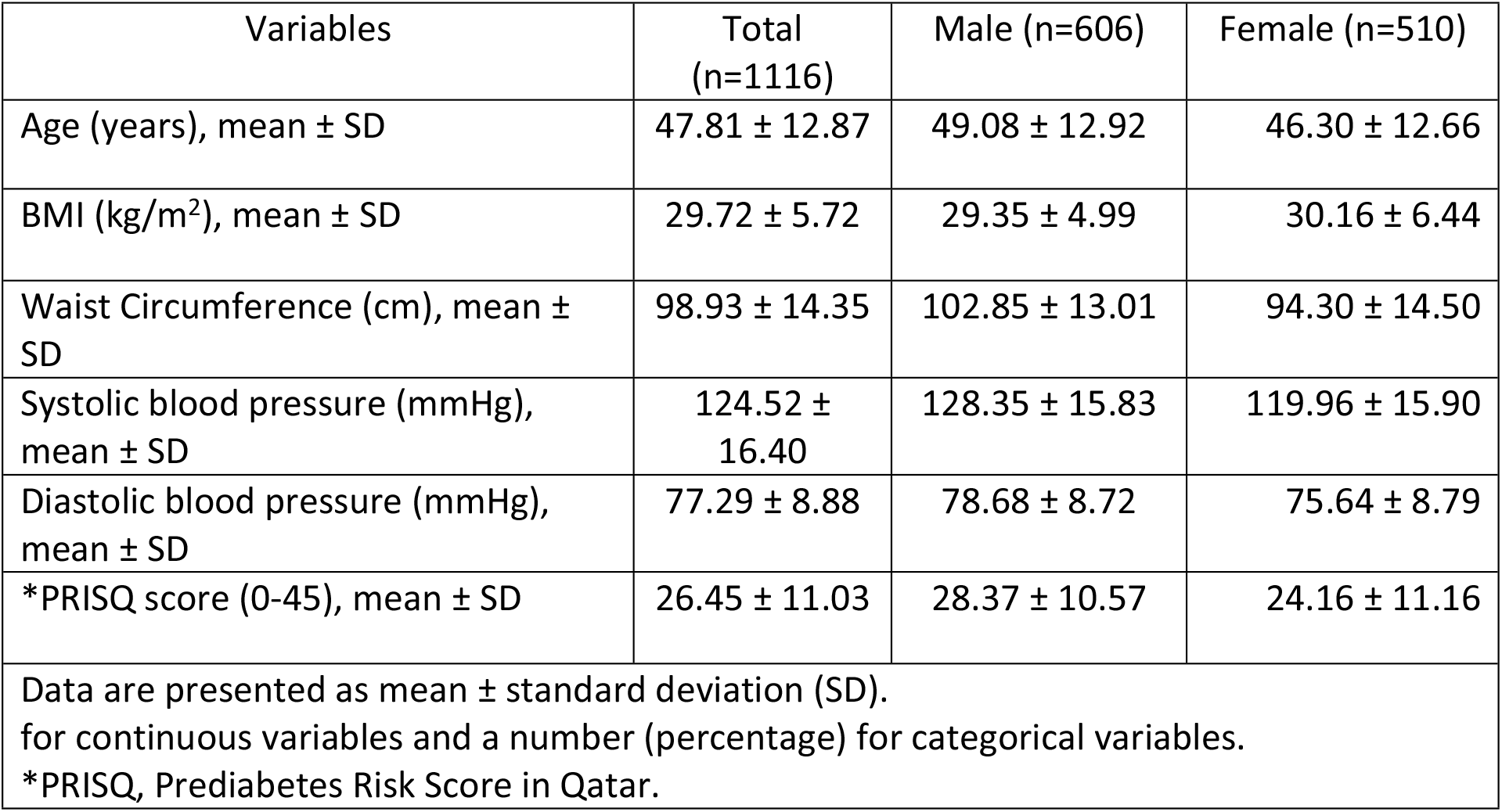
Baseline Characteristics of the Study Population, Stratified by Sex.

| Variables | Total (n=1116) | Male (n=606) | Female (n=510) |
| --- | --- | --- | --- |
| Age (years), mean ± SD | 47.81 ± 12.87 | 49.08 ± 12.92 | 46.30 ± 12.66 |
| BMI (kg/m<sup>2</sup>), mean ± SD | 29.72 ± 5.72 | 29.35 ± 4.99 | 30.16 ± 6.44 |
| Waist Circumference (cm), mean ± SD | 98.93 ± 14.35 | 102.85 ± 13.01 | 94.30 ± 14.50 |
| Systolic blood pressure (mmHg), mean ± SD | 124.52 ± 16.40 | 128.35 ± 15.83 | 119.96 ± 15.90 |
| Diastolic blood pressure (mmHg), mean ± SD | 77.29 ± 8.88 | 78.68 ± 8.72 | 75.64 ± 8.79 |
| *PRISQ score (0-45), mean ± SD | 26.45 ± 11.03 | 28.37 ± 10.57 | 24.16 ± 11.16 |
| Data are presented as mean ± standard deviation (SD).<br>for continuous variables and a number (percentage) for categorical variables.<br>*PRISQ, Prediabetes Risk Score in Qatar. | | | |

### Distribution of PRISQ Scores and Risk Categories

The mean Prediabetes Risk Score Questionnaire (PRISQ) score among the study participants was 26.45 ± 11.03, indicating an overall moderate-to-high prediabetes risk profile within the cohort (Table S1). When stratified according to established PRISQ risk thresholds, nearly half of the participants were classified as high risk for prediabetes (score >27), accounting for 47.7% of the sample (*n* = 532). A further 34.4% (*n* = 384) were categorized as moderate risk (score 17–27), while 17.9% (*n* = 200) were classified as low risk (score ≤16).

Figure 1 illustrates the distribution of participants across PRISQ risk categories, demonstrating a predominance of individuals within the moderate and high-risk groups.

**Figure 1.**
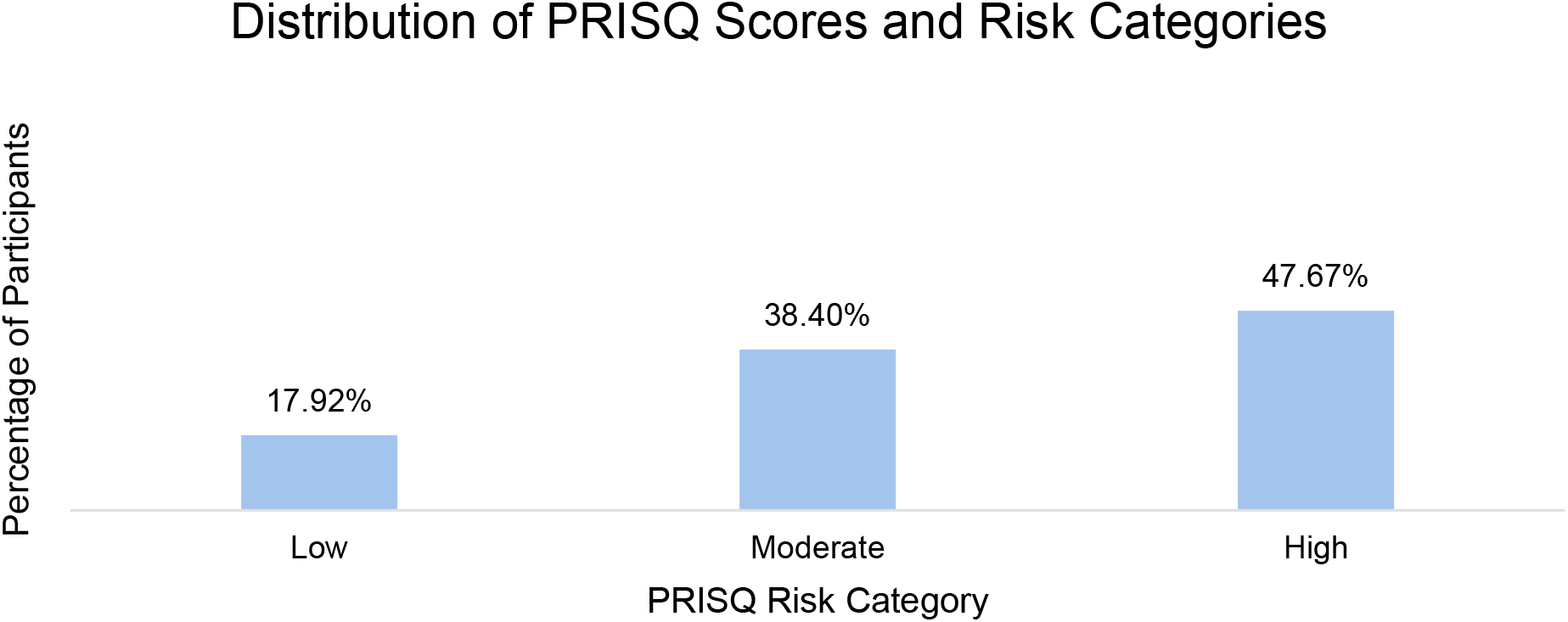
Distribution of PRISQ scores and risk categories.

Age, body mass index (BMI), waist circumference, and blood pressure demonstrated a clear and progressive increase across PRISQ risk categories. Trend analyses confirmed statistically significant differences across low-, moderate-, and high-risk groups for all examined variables (one-way ANOVA, *p* < 0.001 for all). Participants classified within the high-risk PRISQ category were substantially older, with a mean age of 56.2 ± 9.9 years, and exhibited higher anthropometric measures and elevated systolic and diastolic blood pressure values compared with those in the low-risk group. Detailed comparisons of clinical characteristics across PRISQ risk categories are presented in Table 3.

**Table 3.**
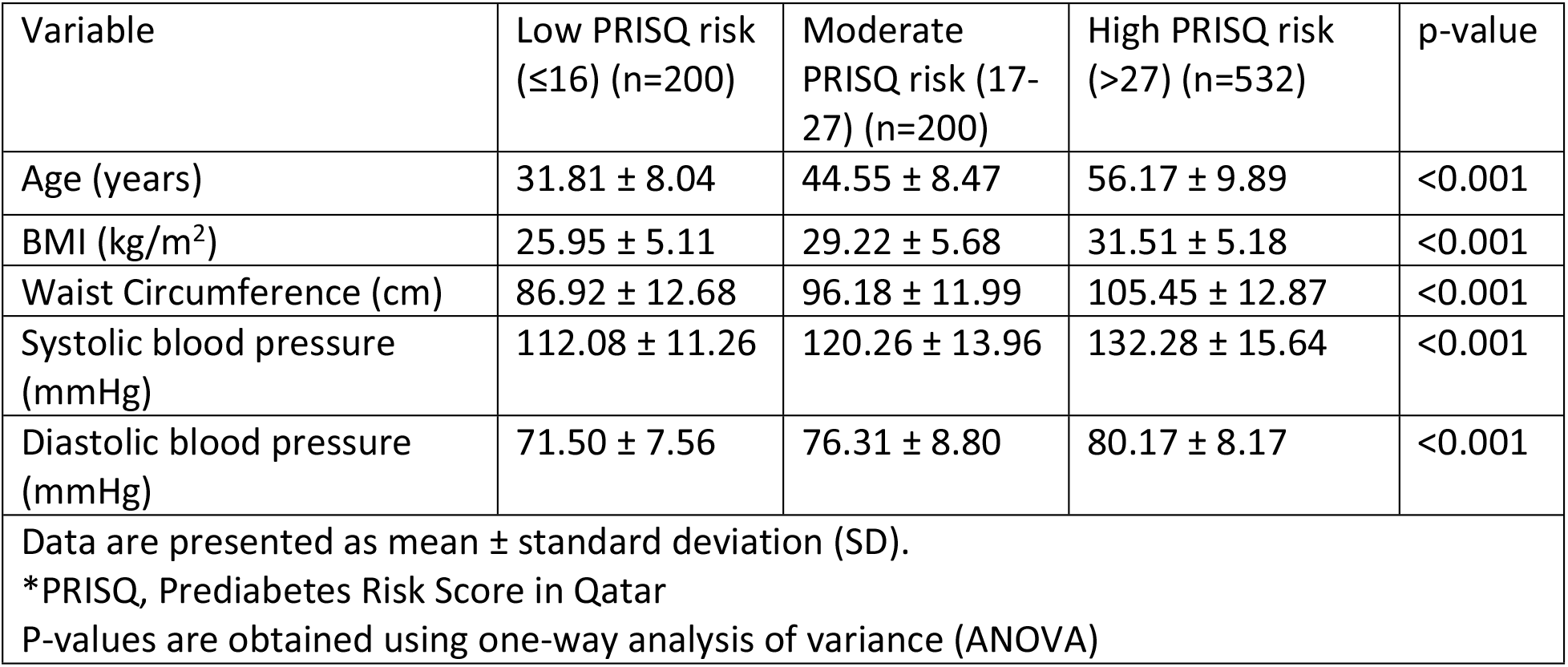
Clinical Characteristics Across PRISQ Risk Categories.

| Variable | Low PRISQ risk ($\leq 16$) (n=200) | Moderate PRISQ risk (17-27) (n=200) | High PRISQ risk ($>27$) (n=532) | p-value |
| --- | --- | --- | --- | --- |
| Age (years) | 31.81 ± 8.04 | 44.55 ± 8.47 | 56.17 ± 9.89 | <0.001 |
| BMI (kg/m<sup>2</sup>) | 25.95 ± 5.11 | 29.22 ± 5.68 | 31.51 ± 5.18 | <0.001 |
| Waist Circumference (cm) | 86.92 ± 12.68 | 96.18 ± 11.99 | 105.45 ± 12.87 | <0.001 |
| Systolic blood pressure (mmHg) | 112.08 ± 11.26 | 120.26 ± 13.96 | 132.28 ± 15.64 | <0.001 |
| Diastolic blood pressure (mmHg) | 71.50 ± 7.56 | 76.31 ± 8.80 | 80.17 ± 8.17 | <0.001 |
| Data are presented as mean ± standard deviation (SD).<br>*PRISQ, Prediabetes Risk Score in Qatar<br>P-values are obtained using one-way analysis of variance (ANOVA) | | | | |

### Factors Associated with PRISQ Score

In multivariable linear regression analysis, the model explained a substantial proportion of variance in PRISQ scores (R^2^ = 0.823, *p* < 0.001). Age demonstrated the strongest positive association with PRISQ score (standardized β = 0.541, *p* < 0.001). Other clinical parameters, including body mass index (BMI; β = 0.270), waist circumference (β = 0.222), systolic blood pressure (β = 0.091), and diastolic blood pressure (β = 0.173), were also independently and positively associated with PRISQ scores (all *p* < 0.001).

Sex was not a statistically significant independent predictor of PRISQ score in the adjusted model (β = −0.273, *p* = 0.40), indicating that male sex did not independently contribute to variation in PRISQ scores within this study population. Full regression results are presented in Table 4.

**Table 4.**
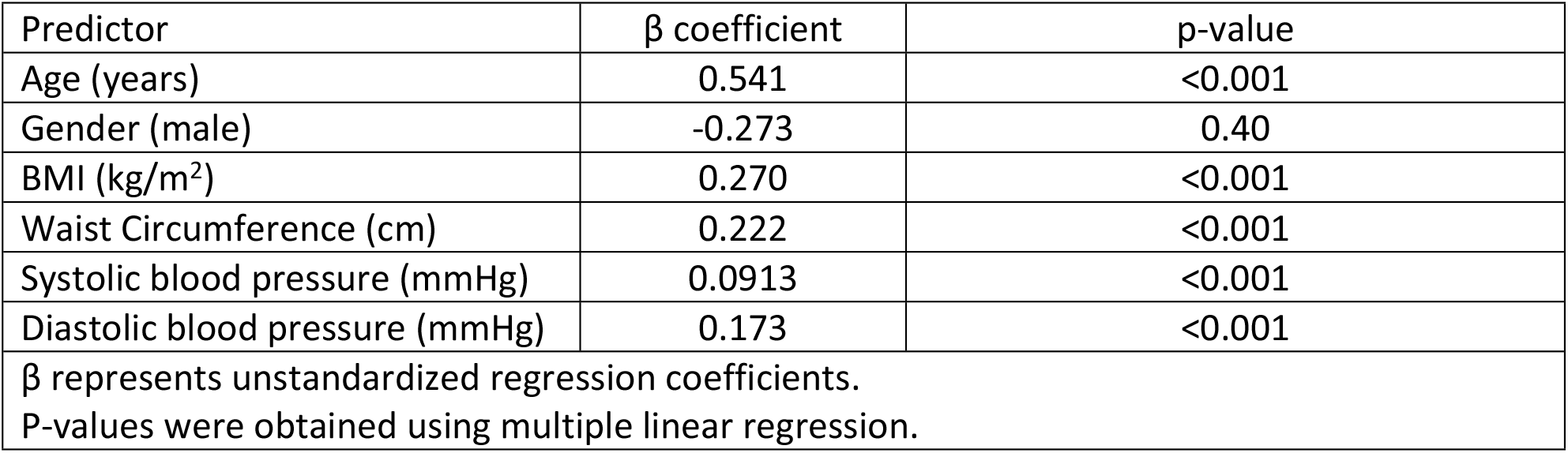
Multiple Linear Regression Analysis of Factors Associated with PRISQ Score.

The distribution of PRISQ risk categories differed significantly by gender (χ^2^ test, *p* < 0.05), as shown in Table S2. A substantially higher proportion of male participants were classified as high risk for prediabetes (55.4%) compared with female participants (38.4%). In contrast, females were more frequently represented in the low-risk category, with 23.9% classified as low risk, compared with 12.9% of males.

Figure 2 illustrates the gender-specific distribution of participants across PRISQ risk categories, highlighting the predominance of high-risk classification among males and a comparatively greater proportion of low-risk classification among females.

**Figure 2.**
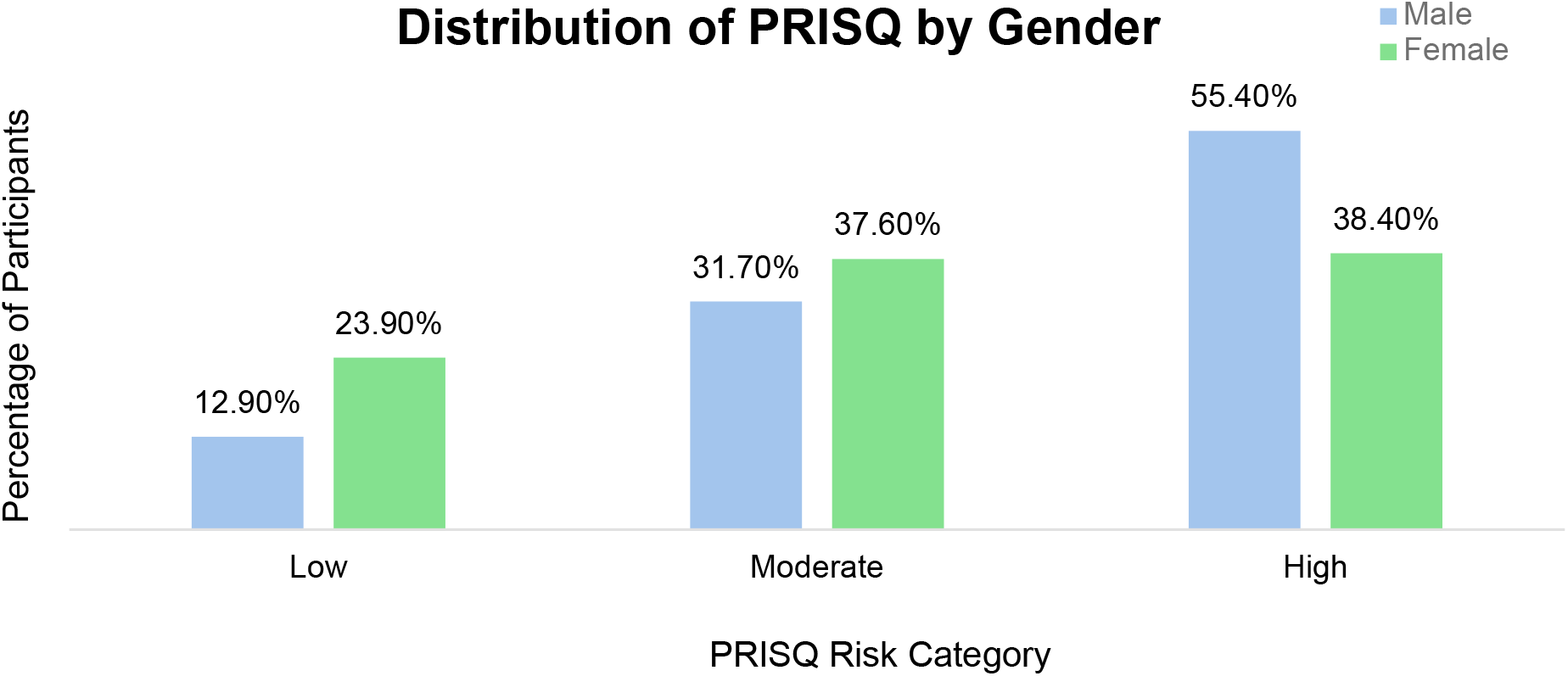
Distribution of PRISQ by gender.

### Sociodemographic Characteristics of the High-Risk PRISQ Population

Among the 532 participants classified as high risk for prediabetes, the largest proportions were observed in the older age groups, with 28.8% aged 50–59 years and 24.6% aged 60 years or older. Most high-risk participants were male, accounting for 63.2% of this subgroup.

In terms of nationality, Northern African (29.1%) and Western Asian (25.4%) individuals constituted the most represented groups within the high-risk category, while Qatari nationals comprised 9.6% of high-risk participants. Most individuals in the high-risk group were either overweight or obese (86.4%).

Educational attainment among high-risk participants was generally high, with more than half (54.1%) reporting completion of a diploma or bachelor’s degree. Most participants (73.3%) had resided in Qatar for more than 10 years. Self-reported physical activity levels of less than 150 minutes per week did not differ significantly within the high-risk group (*p* = 0.097). Detailed sociodemographic characteristics of the high-risk PRISQ population are presented in Table 5.

**Table 5.** Sociodemographic Characteristics of the High-risk PRISQ Population.

| **High Risk Population (n=532)** | **n** | **%** |
| --- | --- | --- |
| **Age (years)** |  |  |
| 18-29 | 34 | 6.4 |
| 30-39 | 74 | 13.9 |
| 40-49 | 140 | 26.3 |
| 50-59 | 153 | 28.8 |
| 60+ | 131 | 24.6 |
| **Gender** |  |  |
| Female | 196 | 36.8 |
| Male | 336 | 63.2 |
| **Nationality** |  |  |
| Qatari | 51 | 9.6 |
| Northern Africa | 155 | 29.1 |
| South-eastern Asia | 63 | 11.8 |
| Southern Asia | 57 | 10.7 |
| Western Asia | 135 | 25.4 |
| Other | 71 | 13.3 |
| **BMI (kg/m<sup>2</sup>)** |  |  |
| Average (<25) | 73 | 13.7 |
| Overweight (25-29.9) | 192 | 36.1 |
| Class 1 Obesity (30-34.9) | 172 | 32.3 |
| Class 2 Obesity (35-39.9) | 68 | 12.8 |
| Class 3 Obesity (40+) | 27 | 5.1 |
| **Educational status** |  |  |
| Never attended school | 6 | 1.1 |
| Primary school (Class 1 to 6) | 20 | 3.8 |
| Secondary school (Class 7 to 12) | 88 | 16.5 |
| Trade/technical/vocational qualification | 42 | 7.9 |
| Diploma/bachelor's degree | 288 | 54.1 |
| Postgraduate degree | 88 | 16.5 |
| **Years of residence in Qatar** |  |  |
| <1 year | 3 | 0.6 |
| 1 to 5 years | 20 | 3.8 |
| > 5 to 10 years | 80 | 15.0 |
| > 10 years | 390 | 73.3 |
| No response | 39 | 7.3 |
| **Average monthly household income (QAR)** |  |  |
| Less than 5000 | 63 | 11.8 |
| 5000 - 10000 | 121 | 22.7 |
| 11000 - 20000 | 120 | 22.6 |
| 21000 - 30000 | 34 | 7.3 |
| 31000 - 40000 | 28 | 5.3 |
| 41000 – 50000 | 12 | 2.3 |
| 51000 - 60000 | 10 | 1.9 |
| More than 60000 | 14 | 2.6 |
| No response | 130 | 24.4 |
| **Physical activity (>150 min/week)** |  |  |
| No | 295 | 55.5 |
| Yes | 237 | 44.5 |
| Data are presented as numbers and percentages. Percentages are calculated within the high-risk PRISQ population (n=532). P-values were obtained using Chi-square goodness of fit tests assuming equal distribution across categories. |  |  |

## Discussion

### Principal Findings of the Study

This study demonstrates a substantial burden of predicted prediabetes risk among adults attending primary care in Qatar, with nearly half of participants (47.7%) classified as high risk using the Prediabetes Risk Score in Qatar (PRISQ). The mean PRISQ score (26.5 ± 11.0) indicates a population skewed toward moderate-to-high risk, driven primarily by older age, higher body mass index, increased waist circumference, and elevated blood pressure, all of which showed strong and statistically significant associations with PRISQ scores in multivariable analysis (R^2^ = 0.823, p < 0.001). These findings directly address the study’s primary objective by confirming that PRISQ identifies a large pool of adults at elevated metabolic risk within the primary care setting who may not yet meet laboratory diagnostic thresholds for dysglycaemia. The marked contrast between the proportion classified as high risk in this study and previously reported national prediabetes prevalence underscores the value of non-invasive, risk-based screening tools in detecting upstream cardiometabolic vulnerability before clinical disease manifests.

### Comparison of key findings with existing Literature

When contextualized within existing literature, the observed risk distribution is consistent with regional evidence indicating a rising burden of cardiometabolic risk across Gulf Cooperation Council populations undergoing rapid urbanization and lifestyle transition. The elevated PRISQ scores observed in this cohort reflect ongoing epidemiological transitions in Qatar. Rapid urbanization has driven substantial lifestyle and dietary changes including reduced physical activity, increased intake of calorie-dense foods, and rising obesity and hypertension which are collectively captured by higher PRISQ scores. These findings align with regional projections from the International Diabetes Federation, which estimate that one in six adults in the Middle East and North Africa (MENA) region currently lives with diabetes and that the burden of prediabetes is expected to rise sharply by 2050[22]. Collectively, these results highlight an urgent need for systematic population-level risk stratification within PHCC to enable early identification and timely intervention.

A major proportion of male population (55.4%) were ranked high-risk in comparison to females (38.4%), underscoring the gender difference. Despite these findings, this association as supported statistically, suggesting that gender does not impact PRISQ score differences. Another important aspect of these findings is that these gender differences are not biologically exerted, other correlated risk factors such as increased waist circumference, BMI, and blood pressure are the mediators explaining increased risk prevalence observed in the male participants. More than gender, these outcomes are likely the results of lifestyle choices which often lead to adverse clinical outcomes. It is recommended that the modifiable risk factors must be accounted especially for males while designing public interventions.

Our findings also pointed towards the significant impact of expatriates and their origins (Western Asian and North African) on the risk score predictions. To contextualize this aspect of finding, a multifactorial point of view is adopted. This risk is characterized by increased prevalence of diabetes and cardiometabolic factors in the origin countries of these expatriates including Egypt, India, and Pakistan, which might explain the higher risk [23, 24]. Then occupational exposure may also play its role since a majority of expatriates are employed in public sector jobs which are often taxing, with longer shift times, further complicated by restricted healthcare access, settling for inexpensive, calorie-dense diets [25]. Another emerging phenomenon is the acculturation of food where expatriates are required to adapt local food customs that include the use of refined carbohydrates, fried foods, and saturated fats, which they might not be accustomed to [26]. Given that middle-aged man are at higher-risk, the lack of cooking facilities due to shared apartments arrangements also contribute in consuming processed or fast food [27]. All the previously mentioned elements are interlinked which demand culturally and occupationally sensitive interventions to provide better health and screening equity to the PHCC attending population.

Across the high-risk population, majority were either overweight or obese individuals, however a notable finding was that individuals in Class III obesity (BMI ≥40 kg/m^2^) constituted a minor proportion. These findings align with other international and regional studies where the burden was exerted more by overweight and Class I/II obesity individuals instead of severely obese individuals [28]. Another possible explanation of Class III obesity being minorly represented in the sample is that the burden of comorbid health conditions is more severe in this category which not just increase the healthcare utilization but requires attending secondary or tertiary care settings instead of primary care (e.g., PHCC) [29]. These findings highlight that more clinical attention should be given to overweight and moderately obese populations, which has elevated representation in high-risk populations.

A notable finding emerged where extended period of stay in Qatar represented a higher risk prediction which suggests the impact is the result of lifestyle acculturation. Expatriates are at increased risk of metabolic disorders given their adoption to local Gulf food and limited physical activity [26]. Similarly, financial conditions also play a significant role. While higher income increases access to healthcare and healthier food choices, their occupational choices encompass office-based environment which are often sedentary [30, 31]. Comparatively, lower income category faces restricted healthcare access, or nutrient-rich food [25]. Emerging evidence suggests that the psychosocial stress of acculturation and socioeconomic transition can further exacerbate cardiometabolic risk among expatriates [32]. These insights position duration of residence and income not as mere background variables but as active social determinants of health, underscoring the need for tailored interventions that address the diverse socioeconomic realities within Qatar’s expatriate population. Collectively, these findings position PRISQ as a pragmatic population-level risk stratification and engagement tool within primary care, with clear relevance for informing targeted prevention, health education, and early intervention strategies in Qatar’s diverse and aging population.

### Strengths and Limitations of the Study

This study has several notable strengths. It draws on data from the HEALTHSIGHT study,[21] which employed a stratified random sampling strategy across age, sex, and nationality, enhancing the representativeness of adults registered with the Primary Health Care Corporation (PHCC) in Qatar. Use of a nationally distributed primary care network enabled assessment of prediabetes risk in a real-world healthcare setting, directly relevant to population-level prevention strategies. The study applied PRISQ, a validated, non-invasive risk score specifically developed for the Qatari population using routinely collected clinical parameters, strengthening internal validity and local applicability. Integration of objectively measured data from electronic medical records alongside standardized anthropometric assessments reduced reliance on laboratory testing and supports feasibility for large-scale implementation in primary care. The availability of comprehensive sociodemographic and clinical information enabled detailed subgroup analyses by sex, age, and nationality, facilitating identification of population groups with disproportionate risk. As a HEALTHSIGHT-derived analysis, the study further benefits from standardized data collection protocols, ethical oversight, and harmonization with national health research initiatives, enhancing comparability with related studies.

The study has several limitations which should be acknowledged. The cross-sectional design limits causal inference and precludes assessment of temporal changes in prediabetes risk or progression to diabetes, such that findings represent risk distribution at a single time point. Although the sampling strategy aimed to be representative, the final analytic sample constituted a subset of the HEALTHSIGHT cohort due to missing data and non-response, introducing potential selection bias toward individuals more engaged with primary care; caution is therefore warranted when generalizing beyond PHCC-registered adults with available electronic health records. Use of nationality as a proxy for ethnicity may mask heterogeneity within broad population groups and limit generalizability, while self-reported lifestyle measures, including physical activity, are subject to recall and reporting bias despite standardized procedures. Although PRISQ is validated in its derivation cohort, it does not explicitly capture region-specific behavioral and environmental factors such as dietary patterns, occupational constraints, cultural practices, or vitamin D status which may influence metabolic risk and are only indirectly reflected through clinical parameters. The absence of longitudinal follow-up and outcome ascertainment further limits evaluation of progression to incident diabetes, underscoring the need for prospective validation within PHCC or linkage to national registries.

### Policy Implications and Suggestions for Further Research

The high prevalence of elevated prediabetes risk identified in this study has important implications for public health policy and preventive care delivery in Qatar. Integrating PRISQ into routine Primary Health Care Corporation (PHCC) workflows could enable systematic, non-invasive risk stratification in primary care and support earlier identification of individuals who may benefit from targeted preventive interventions before progression to overt diabetes. As a quantified risk score, PRISQ may also serve as a practical communication tool during clinical encounters, facilitating shared decision-making by translating abstract metabolic risk into a tangible metric and demonstrating the potential impact of modifiable behaviors, including weight management, dietary improvement, physical activity, and blood pressure control. This approach may be particularly valuable for younger adults who do not yet meet diagnostic thresholds for dysglycaemia but already exhibit adverse cardiometabolic profiles. At a systems level, embedding PRISQ within electronic health record–supported workflows could assist PHCC in prioritizing preventive services, optimizing resource allocation, and aligning screening strategies with national non-communicable disease prevention goals. Given the mediating role of modifiable factors such as adiposity and blood pressure observed in this study, population-level lifestyle interventions are likely to yield substantial public health benefits.

Further research should prioritize longitudinal follow-up of individuals classified as high risk by PRISQ to assess progression to prediabetes, type 2 diabetes, and cardiovascular outcomes, and to evaluate predictive performance over time in the Qatari primary care context. Once longitudinal validity is established, serial PRISQ assessments could be explored for monitoring risk trajectories and reinforcing sustained lifestyle modification. Additional work incorporating context-specific behavioral, occupational, and environmental determinants—particularly within Qatar’s diverse expatriate workforce—alongside qualitative research on barriers to prevention, would further strengthen the evidence base for implementation.

## Conclusion

This study demonstrates a substantial burden of predicted prediabetes risk among adults attending primary care in Qatar, with nearly half classified as high risk using the PRISQ tool. Application of a population-specific, non-invasive risk score in a real-world Primary Health Care Corporation (PHCC) setting provides insight into cardiometabolic risk distribution across a diverse population. Older age, greater adiposity, elevated blood pressure, male predominance within high-risk strata, and longer residence in Qatar emerged as key contributors, reflecting the interaction of sociodemographic and metabolic determinants in a rapidly urbanizing context.

The findings support PRISQ as a risk stratification and engagement tool rather than a diagnostic instrument. Translating cardiometabolic risk into a simple score may facilitate shared decision-making in primary care, particularly for younger adults with adverse risk profiles but without overt dysglycaemia. Integration into routine PHCC workflows could strengthen early identification and targeted prevention; however, longitudinal validation is required before PRISQ can inform risk trajectories or follow-up strategies. Aligning risk-based screening with culturally appropriate preventive interventions may contribute meaningfully to national efforts to reduce the burden of diabetes and cardiometabolic disease.

## Data Availability

all the data is already presented in the manuscript.

## Declarations

## Abbreviations

BMI: Body mass index
T2DM: Type 2 Diabetes Mellitus
EMR: Electronic medical record
PHCC: Primary Health Care Corporation

## Ethics approval and consent to participate

All study procedures were conducted in accordance with the Declaration of Helsinki. The study was reviewed and approved by Primary Health Care Corporation (PHCC)’s Independent Review Board (BUHOOTH-D-23-00058). Informed consent was obtained from participants aged 18 or over. Overall, the study was planned and conducted with integrity according to generally accepted ethical principles.

## Consent for publication

Not applicable

## Availability of data and materials

All data generated or analyzed during this study are included in this published article and its supplementary information files.

## Authors contributions

1. Dana Bilal El Kaissi: Conception of idea, Literature review, drafting of manuscript, data analysis and interpretation and review of manuscript
2. Mohamed Ahmed Syed: Conception of idea, literature review, data analysis and interpretation drafting and review of manuscript
3. Muslim Abbas Syed: Literature review, drafting of manuscript, data analysis and interpretation and drafting and review of manuscript

## Acknowledgements

We would like to acknowledge Primary Health Care Corporation for funding this study and the data collectors who were involved in data collection.

## Clinical trial number

not applicable

